# Predictors of Brain Injury: The Performance of Biomechanical Head Acceleration Severity Metrics for Concussion Prediction in Men’s and Women’s Rugby League Players

**DOI:** 10.64898/2026.08.12.26360260

**Authors:** James Tooby, Cameron Owen, Sarah Whitehead, Sean Scantlebury, Dane Vishnubala, Lyndia Wu, Matthew Kitchin, Songbai Ji, Steve Rowson, Ross Tucker, Chaokai Zhang, Ben Jones

## Abstract

**Objective:** Describe and compare the biomechanical severity of head acceleration events (HAEs) associated with diagnosed concussion in elite men’s and women’s rugby league using instrumented mouthguards (iMGs) and evaluate the diagnostic accuracy and screening performance of severity metrics within the current Head Injury Assessment (HIA) process.

**Methods:** A prospective cohort of 398 men and 252 women from Super League teams wore iMGs across 515 matches. Following a matching and data quality screening procedure, 123 HIAs (106 men, 17 women) were matched to HAEs, 52 of which were diagnosed concussions (44 men, 8 women). High-magnitude asymptomatic control HAEs (22,676 men, 3,200 women) were sampled proportionally to the number of HIAs. A range of biomechanical severity metrics were calculated for HAEs. Statistical comparisons between outcomes were made. Receiver operating characteristic (ROC) analysis evaluated diagnostic accuracy (ability to predict diagnosed concussions within the HIA). Precision-recall analysis evaluated screening performance (ability to discriminate observable concussion signs from asymptomatic events).

**Results:** Diagnosed concussions had greater severity than asymptomatic HAEs across all metrics in both sexes. For diagnostic accuracy, area under the ROC curve ranged from 0.61 to 0.72 in men and 0.53 to 0.86 in women. For screening performance, optimal thresholds in several metrics provided theoretical improvements to precision over current thresholds used in rugby, but recall remained <0.20.

**Conclusion:** These findings support integrating iMG-derived severity metrics into a multimodal, clinician-led HIA pathway as objective adjuncts for diagnosis and screening, while reinforcing that they cannot replace clinical judgement or other assessment modalities.

**What is already known on this topic:** Instrumented mouthguards are increasingly used in rugby to quantify head acceleration events and trigger Head Injury Assessment (HIA) alerts, but current screening thresholds based on simple peak kinematics have low sensitivity for identifying HAEs linked with visible concussion signs, and very few iMG-measured concussions, particularly in women, have been reported in current research.

**What this study adds:** This study provides the largest dataset of iMG-measured concussions in any sport, shows that concussive HAEs are more severe than asymptomatic events across multiple biomechanical metrics in both sexes, and identifies several severity metrics with diagnostic accuracy comparable to existing HIA sub-tests and modest theoretical improvements over current screening thresholds.

**How this study might affect research, practice or policy:** These findings support incorporating iMG-derived severity metrics as objective adjuncts within clinician-led HIA pathways, highlight the need for sex-inclusive iMG datasets and multimodal concussion identification, and may inform future refinement of iMG screening thresholds in elite rugby and other contact sports.

## Introduction

Sports-related concussions constitute approximately 20% of all traumatic brain injuries (TBIs) within the general population^1^ and are amongst the most common injuries in contact sports^2^. Post-concussion symptoms, including cognitive and sensorimotor impairment, can persist for weeks to months^3,4^, and multiple concussions are associated with long-term cognitive deficits^5^. Consequently, the prevention, detection, and management of concussion are a priority in contact sports.

Consensus defines sport-related concussion as TBI induced by biomechanical forces^6^, therefore biomechanics has a central role in understanding concussions^7^. Wearable inertial sensors can approximate biomechanical forces and resulting tissue-level responses in TBI mechanisms by capturing short periods of head kinematics, known as head acceleration events (HAEs). Instrumented mouthguards (iMGs) have emerged as a popular sensor due to superior skull coupling compared to other sensor mountings^8^ and are widely used in rugby^9,10^ and other contact sports^11^.

Instrumented mouthguard data have been integrated into rugby’s Head Injury Assessment (HIA) protocol, a multimodal process in which players with potential head injuries are removed from play for off-field clinical review^12,13^. Potential head injuries are identified based on observable concussion signs by recognised by sideline medical practitioners^14^. The recent addition of iMG data to this process entails an alert to if an HAE exceeds a pre-determined severity threshold to initiate an off-field assessment. This is an important initiative, given that 20% of rugby league players reportedly hide concussion symptoms from medical staff^15,16^.

The iMG-based screening alert was introduced pragmatically within rugby. Current thresholds were selected to avoid overloading the HIA process with excessive player removals and rely on simple peak kinematic measures, which may be oversimplified as they do not capture the direction and duration of HAEs. This oversimplification may contribute to the low sensitivity of current thresholds for identifying HAEs that were associated visible signs of concussion in elite-level men’s and women’s rugby union, which was estimated at below 20%^17^. Other established biomechanical severity metrics may offer additional value for approximating TBI mechanisms, including mechanical energy measures^18^, brain strain estimates^19^, and composite injury criteria^20–25^.

Research on severity metrics for clinically diagnosed concussions using iMG data remains limited; a recent meta-analysis of concussions measured by wearable sensors reported only six iMG-measured concussions, and none of these were in women^26^. Accordingly, this study aimed to: describe the severity of HAEs associated with diagnosed concussion over two seasons of elite men’s and women’s rugby league matches using a range of biomechanical metrics; compare the severity of HAEs resulting in diagnosed concussion, observable signs of concussion, and high-magnitude asymptomatic control events, and between sexes; and to assess the predictive capability of each severity metric, including its diagnostic accuracy and screening performance within the current HIA process.

## Methods

### Participants

A prospective observational cohort study was conducted across all teams in the Men’s and Women’s Super League competitions during the 2024 and 2025 seasons, in which 398 men and 252 women wore custom-fit instrumented mouthguards (Prevent Biometrics, Minneapolis, MN, USA) across 515 matches (*n*=354 men’s, 161 women’s). Ethics approval was granted by the Leeds Beckett University Research Ethics Committee (REF: 100411).

### Instrumented Mouthguards

Instrumented mouthguards were configured consistently with previous rugby league research^10,27^. Data processing included two algorithms to classify HAEs as true-or false-positives and as containing low, moderate, or severe noise. All match-day HAEs exceeding 5 *g* at the head centre-of-gravity and 400 rad/s^2^ were accessed for analysis (*n*=333,957 men’s, 81,980 women’s).

### Severity Metrics

Severity metrics were selected from prior research, grouped into four categories: peak kinematics, mechanical energy metrics, composite injury criteria, and brain strain metrics. All metrics were calculated from processed head kinematics time-series data using equations shown in Table 1.

**Table 1.**
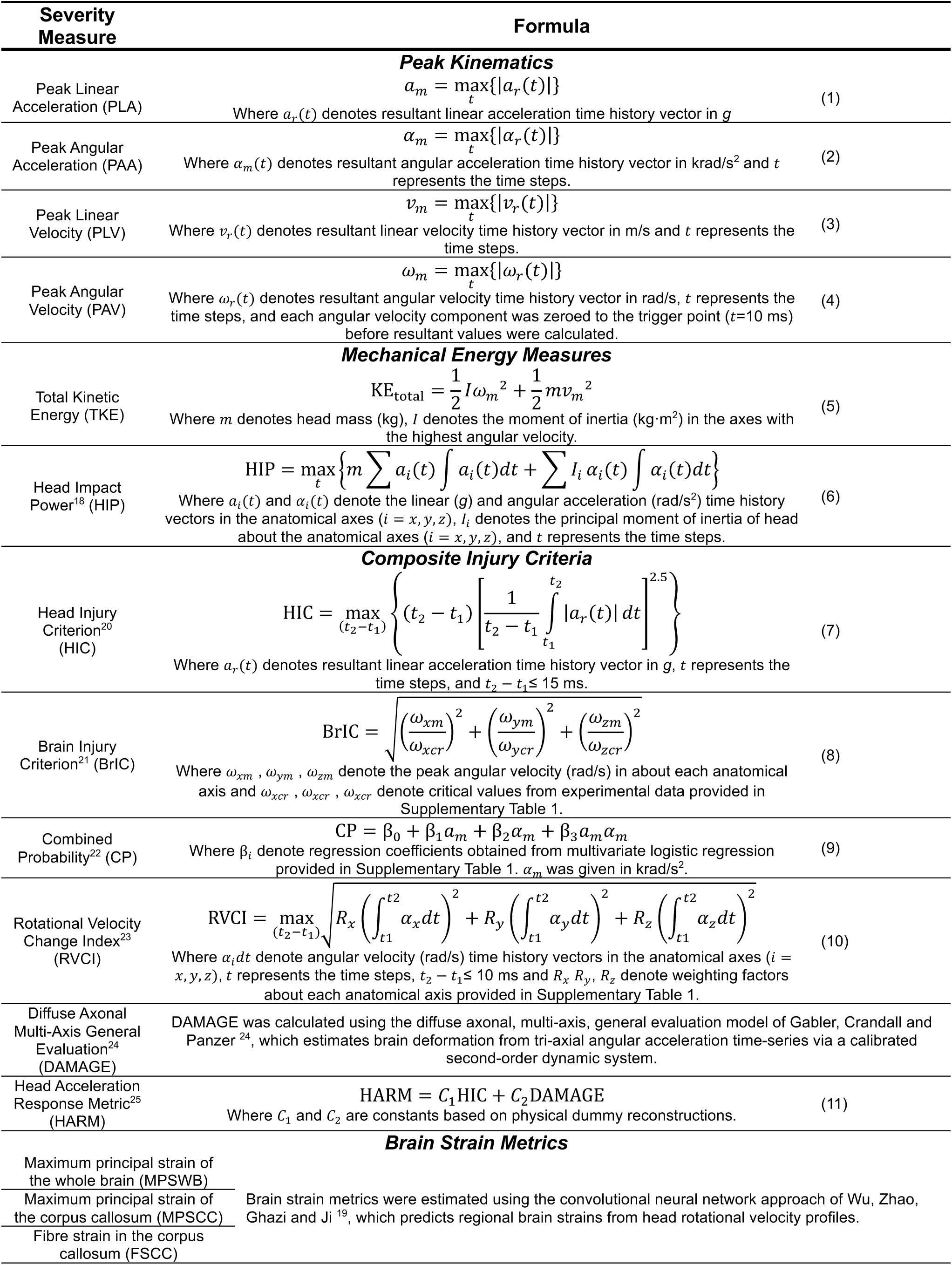
Equations and methods used to calculate peak kinematics, mechanical energy measures, composite injury criteria, and brain strain metrics.

### Head Injury Assessment Protocol

The HIA protocol was implemented across all matches, whereby observable concussion signs were identified with the use of video-replays and players were removed from play for off-field clinical assessment^12,13^. A match-day doctor determined the outcome of each HIA, informed by the Sport Concussion Assessment Tool^28^, as diagnosed concussion (fail) or return to match participation (pass). Data were collected before iMG data were introduced into the HIA, and each HIA was recorded and reported to the Rugby Football League (RFL) as per medical standards practices.

### Matching and Data Quality Screening Procedure

A four-stage hierarchical screening process (Figure 1a) was implemented to retain only HIAs with high-quality kinematic signals, based on: (1) an iMG recording during the match; (2) observable concussion signs following an identifiable causative event (i.e., head acceleration from which signs originated); (3) corresponding HAE for causative event; and (4) high-quality HAE signal (i.e., minimal noise based on Prevent Biometrics algorithm). Stage 2 was performed by a video analyst, independent of iMG data, using consensus definitions of observable concussion signs^14^. Stage 3 cross-referenced video footage of the causative event with time-synchronised iMG data using a graphical user interface (Supplementary Figure 1). Exclusions were made when missing or erroneous iMG data led to an inaccurate approximation of the causative event. Stage 4 excluded HAEs classified as moderate or severe noise.

**Figure 1.**
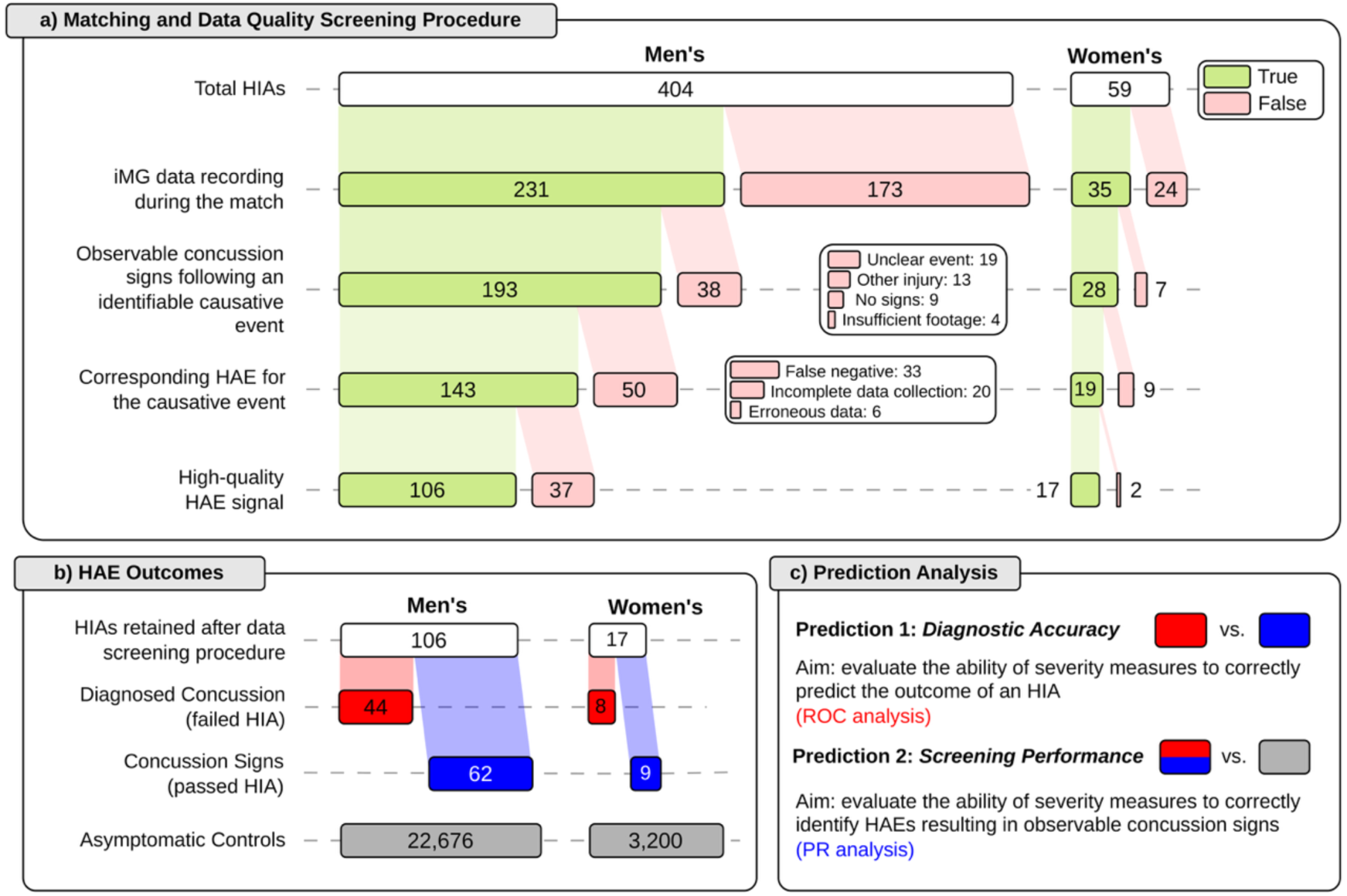
(a) Inclusions and exclusions made during the matching and data quality screening procedure; (b) The number of HAEs for each outcome used within the analysis; (c) Prediction 1, where the positive class was diagnosed concussion and the negative class was observable concussion signs, and Prediction 2, where the positive class was both diagnosed concussions and observable concussion signs and the negative class was asymptomatic controls. HIA = head injury assessment, iMG = instrumented mouthguard, HAE = head acceleration event, ROC = receiver operating characteristic, PR = precision-recall

### High-Magnitude Asymptomatic Control Events

Control events were sampled from HAEs recorded during match-play that were classified as true positives with minimal noise. Only higher-magnitude HAEs were included to isolate the top 25% of HAEs, consistent with previous research^22^, so HAEs exceeding the 87.5^th^ percentile of PLA, PAA, or PAV were retained, resulting in retention of 24.6% and 24.3% of men’s and women’s HAEs, respectively. Percentiles were calculated separately for men (PLA: 20.5 *g*, PAA: 1.7 krad/s^2^, PAV: 15.0 rad/s) and women (PLA: 20.0 *g*, PAA: 1.7 krad/s^2^, PAV: 15.0 rad/s). Potentially symptomatic HAEs were removed by excluding all HAEs recorded during player-matches in which the player was removed for a HIA (*n*=463 player-matches), or when an RFL analyst flagged a *potential* observable concussion sign (*n*=2,323 player-matches), the latter being identified as part of a standard RFL post-match review to inform potential post-match clinical assessment.

Control events were sampled proportionally to the number of HIAs retained after data screening^22^. Across the study period, HIA rates were 32.0 and 9.7 per 1,000 hours for men and women, respectively. Therefore, 106 men’s and 17 women’s HIAs equated to 3,310 and 1,756 player-hours of exposure, respectively, which accounted for approximately 27.0% and 31.5% of all player-hours across the two seasons. This exposure resulted in proportional control sample sizes of 22,676 HAEs in men and 3,200 HAEs in women, based on previous HAE rates^10,27^.

### Statistical Analysis

Generalised linear mixed models (GLMMs) were fitted for each severity metric using the *glmmTMB* package^29^, with outcome, sex, and their interaction as fixed effects and player and team as random intercepts. Outcomes were: diagnosed concussion (i.e., failed HIA), observable concussion signs (i.e., entry to HIA process due to exhibiting observable concussion signs but passed), and asymptomatic controls (Figure 1b). Estimated marginal means and 95% confidence intervals (CIs) were obtained on the response scale, and pairwise comparisons between outcomes were performed within and between sexes via the *emmeans* package using Tukey-adjusted comparisons^30^. Model fit and residual diagnostics were assessed. As the GLMM for CP did not converge, CP was presented descriptively using median and interquartile range (IQR) values, with no statistical comparisons for this severity measure.

Each severity metric was evaluated for two predictions: (1) *diagnostic accuracy* and (2) *screening performance* (Figure 1c). Positive and negative classes were based on HAE outcomes: for prediction 1, the positive class was diagnosed concussions (failed HIA) and the negative class was concussion signs (passed HIA); for prediction 2, the positive class comprised both diagnosed concussion and concussion signs, and the negative class was asymptomatic control HAEs. Receiver operating characteristic (ROC) analysis was conducted for prediction 1, including the calculation of true positive rate (TPR, eq. 12), false positive rate (FPR, eq. 13), and area under the ROC curve (ROC-AUC) and the optimal decisioning threshold was

identified using Youden’s Index (*J*, eq. 15). Precision-recall (PR) analysis was conducted for prediction 2 due to the imbalance between positive and negative classes; precision (eq. 14) and recall (eq. 12) were calculated, and the optimal decisioning threshold was based on F_β_ score (eq. 16). A β value of 0.5 was selected to prioritise precision over recall, reflecting the need to avoid excessive false-positive alerts when using iMGs to initiate the HIA process. F_β_ values for each severity metric were compared to the predictive performance of the current threshold implemented by World Rugby and the RFL for screening players (i.e., PLA ≥75 *g* and PAA ≥4.5 krad/s^2^ for men; PLA ≥55 *g* and PAA ≥4.5 krad/s^2^ for women). The relative performance of severity metrics for each prediction and sex was assessed descriptively by ranking based on ROC-AUC (prediction 1) and F_0.5_ (prediction 2) values. Stratified bootstrap resampling with 1,000 resamples was used to estimate 95% CIs for all ROC and PR values, with resampling performed separately within each outcome class to preserve original class proportions.

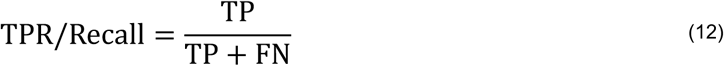

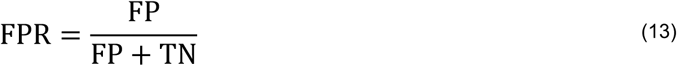

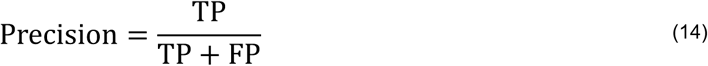

Where TP, TN, FP, FN are the number of true positives, true negatives, false positives, and false negatives, respectively.

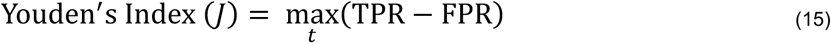

Where *t* represents all threshold values.

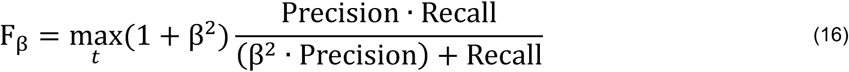

Where β is 0.5 and *t* represents all threshold values.

## Results

Overall, 123 HIAs were included (106 men, 17 women), including 52 diagnosed concussions (44 men, 8 women), recorded across 103 unique players (87 men, 16 women). A further 25,876 asymptomatic control HAEs (22,676 men, 3,200 women) were included from these players and an additional 477 players (262 men, 215 women).

Figure 2 shows the distribution of each severity metric for each HAE outcome in men and women. Diagnosed concussions were significantly greater than asymptomatic controls in all severity metrics for both sexes, except for CP, which was not compared statistically due to lack of model convergence. In men, HAEs with observable concussion signs were also significantly greater than asymptomatic controls in all statistically tested metrics, whereas in women this was observed for only seven severity metrics (PLA, PAA, HIP, HIC, RVCI, HARM). HAEs associated with failed HIAs (i.e., diagnosed concussion) were significantly greater than those with passed HIAs (i.e., observable concussion signs) in nine severity metrics for men (PAV, HIP, BrIC, RVCI, DAMAGE, HARM, MPSWB, MPSCC, FSCC) and three severity metrics for women (BrIC, MPSWB, MPSCC). Significant sex differences were identified for four metrics for diagnosed concussions (HIP, HIC, MPSCC, FSCC) and eight severity metrics for observable concussion signs (PAA, HIP, BrIC, RVCI, HARM, MPSWB, MPSCC, FSCC). Supplementary Table 2 shows mean values and 95% CIs for each outcome and severity metric and Supplementary Tables 3 and 4 show estimated mean differences and *p* values for all pairwise comparisons.

**Figure 2.**
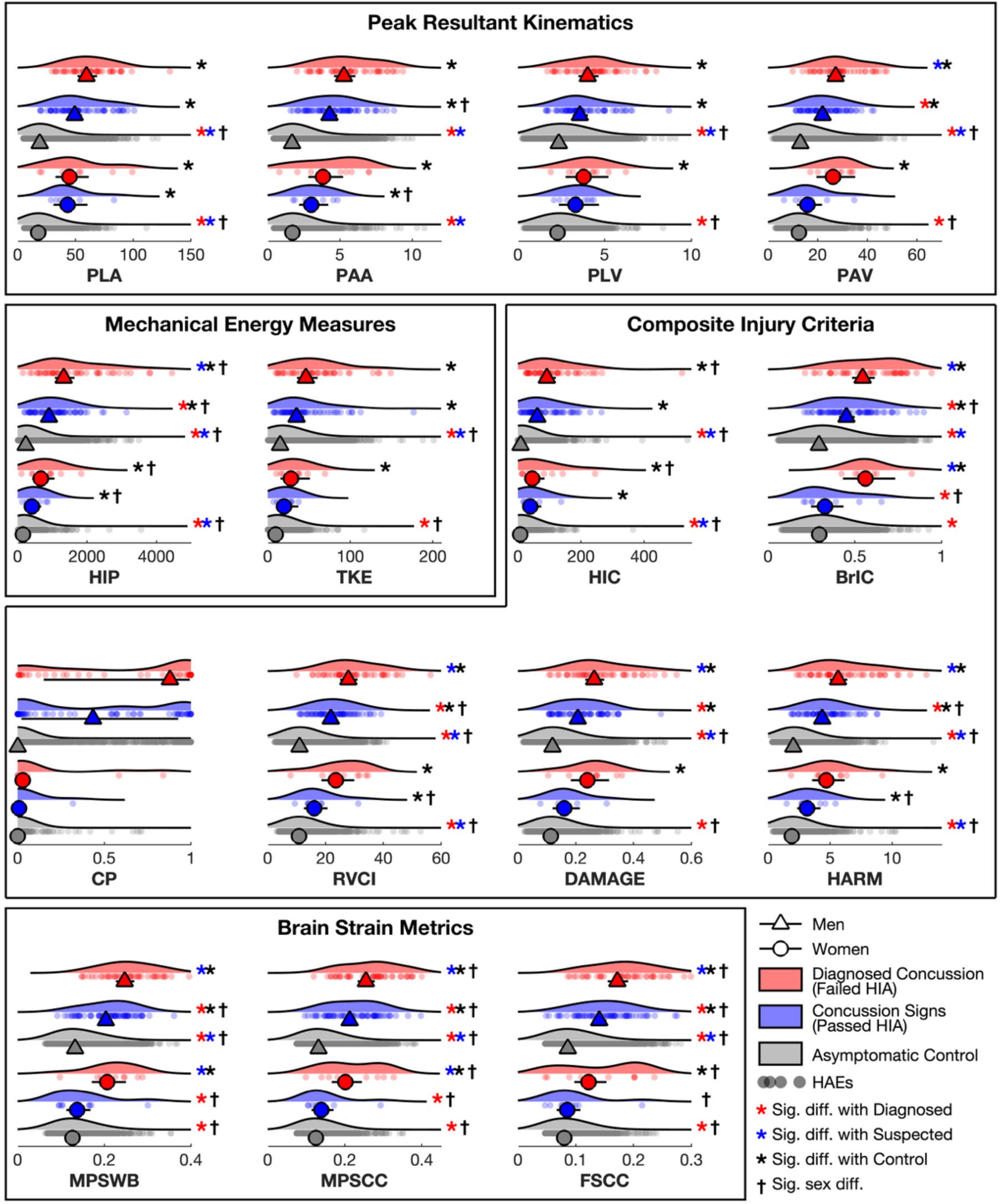
The distribution of severity metrics for men’s and women’s HAEs resulting in diagnosed concussion and observable concussion signs compared with asymptomatic controls. Triangles (men) and circles (women) with whiskers represent mean values and 95% CIs for each outcome. Significant differences (*p* < 0.05) are indicated with asterisks. Median and IQR values are presented for CP due to lack of model convergence. Supplementary Tables 2-4 show tabulated results. PLA = peak linear acceleration, PAA = peak angular acceleration, PLV = peak linear velocity, PAV = peak angular velocity, HIP = Head Impact Power, TKE = total kinetic energy, HIC = Head Injury Criterion, BrIC = Brain Injury Criterion, RVCI = Rotational Velocity Change Index, CP = Combined Probability, DAMAGE = Diffuse Axonal Multi-Axis General Evaluation, HARM = Head Acceleration Response Metric, MPS = maximum principal strain of the whole brain, MPSCC = maximum principal strain of the corpus callosum, FSCC = fibre strain of the corpus callosum, HIA = Head Injury Assessment, HAE = head acceleration event

For prediction 1 (*diagnostic accuracy*), ROC-AUC values ranged from 0.61 to 0.72 in men, with RVCI, MPSWB, and PAV ranking the highest, and 0.53 to 0.86 in women, with BrIC, PAV, and RVCI ranking the highest. For prediction 2 (*screening performance*), current screening thresholds resulted in F_β_ scores of 0.30 (CI:0.20-0.40) in men and 0.18 (CI:0.00-0.41) in women. Precision and recall were 0.35 (CI:0.24-0.47) and 0.18 (CI:0.11-0.25) in men and 0.12 (CI:0.00-0.27) and 0.18 (CI:0.11-0.25) in women. Greater F_β_ scores were achieved by optimal decisioning thresholds in nine severity metrics in men (PLA, PAA, TKE, HIC, CP, RVCI, HARM, MPSWB, MPSCC) and 12 severity metrics in women (PLA, PLV, TKE, HIC, BRIC, CP, RVCI, DAMAGE, HARM, MPSWB, MPSCC, FSCC), although all CIs overlapped (Figure 3). Based on F_β_, the highest-ranking severity metrics were HARM, RVCI, and HIC in men and PLA, FSCC, and RVCI in women.

**Figure 3.**
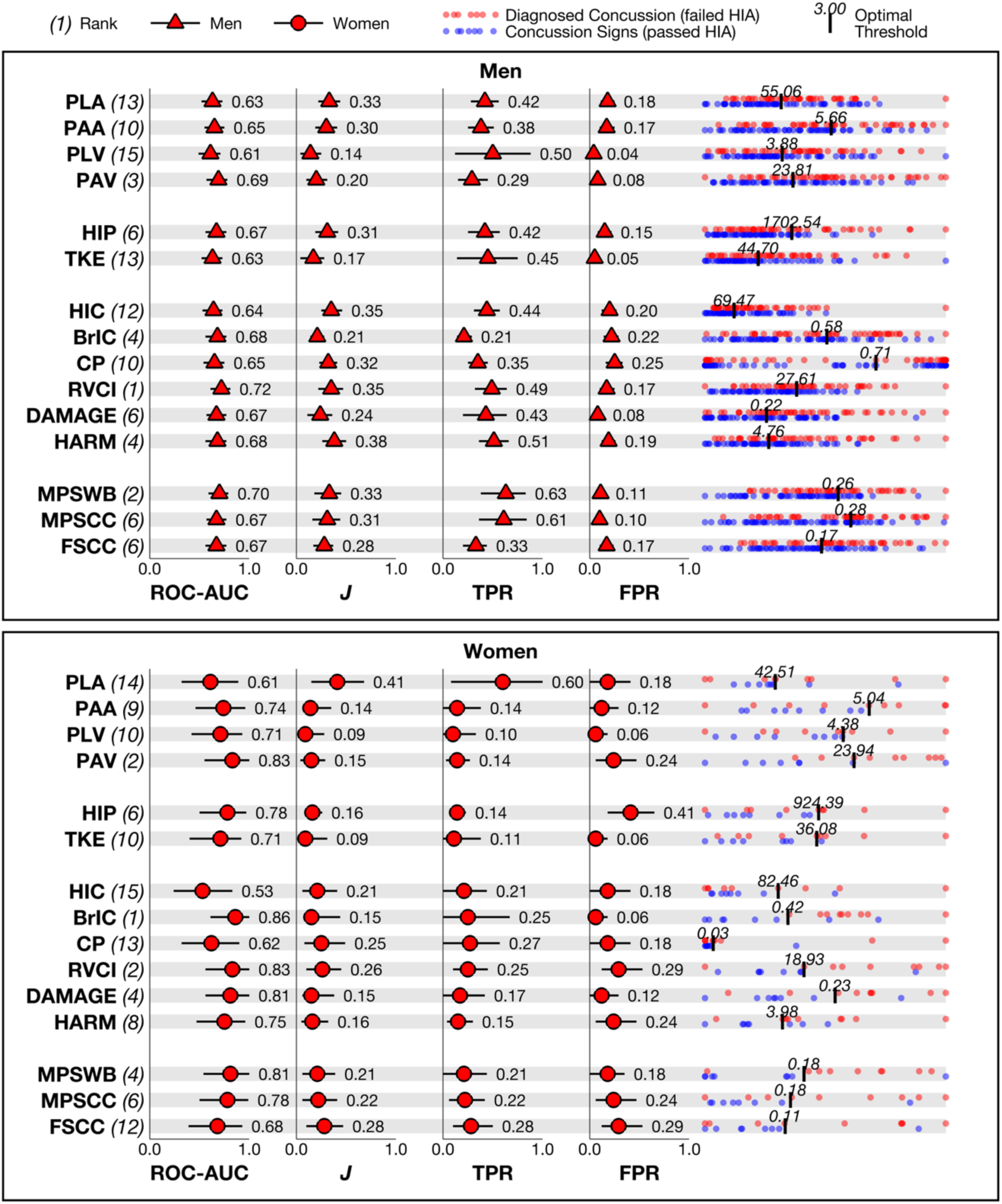
Diagnostic accuracy of each severity metric for men and women based on ROC analysis. Optimal decisioning thresholds were based on maximising *J*. TPR and FPR values correspond with optimal decisioning thresholds. Rank was based on ROC-AUC values. Supplementary Tables 5 shows tabulated results. HIA = Head Injury Assessment, PLA = peak linear acceleration, PAA = peak angular acceleration, PLV = peak linear velocity, PAV = peak angular velocity, HIP = Head Impact Power, TKE = total kinetic energy, HIC = Head Injury Criterion, BrIC = Brain Injury Criterion, RVCI = Rotational Velocity Change Index, CP = Combined Probability, DAMAGE = Diffuse Axonal Multi-Axis General Evaluation, HARM = Head Acceleration Response Metric, MPS = maximum principal strain of the whole brain, MPSCC = maximum principal strain of the corpus callosum, FSCC = fibre strain of the corpus callosum

**Figure 4.**
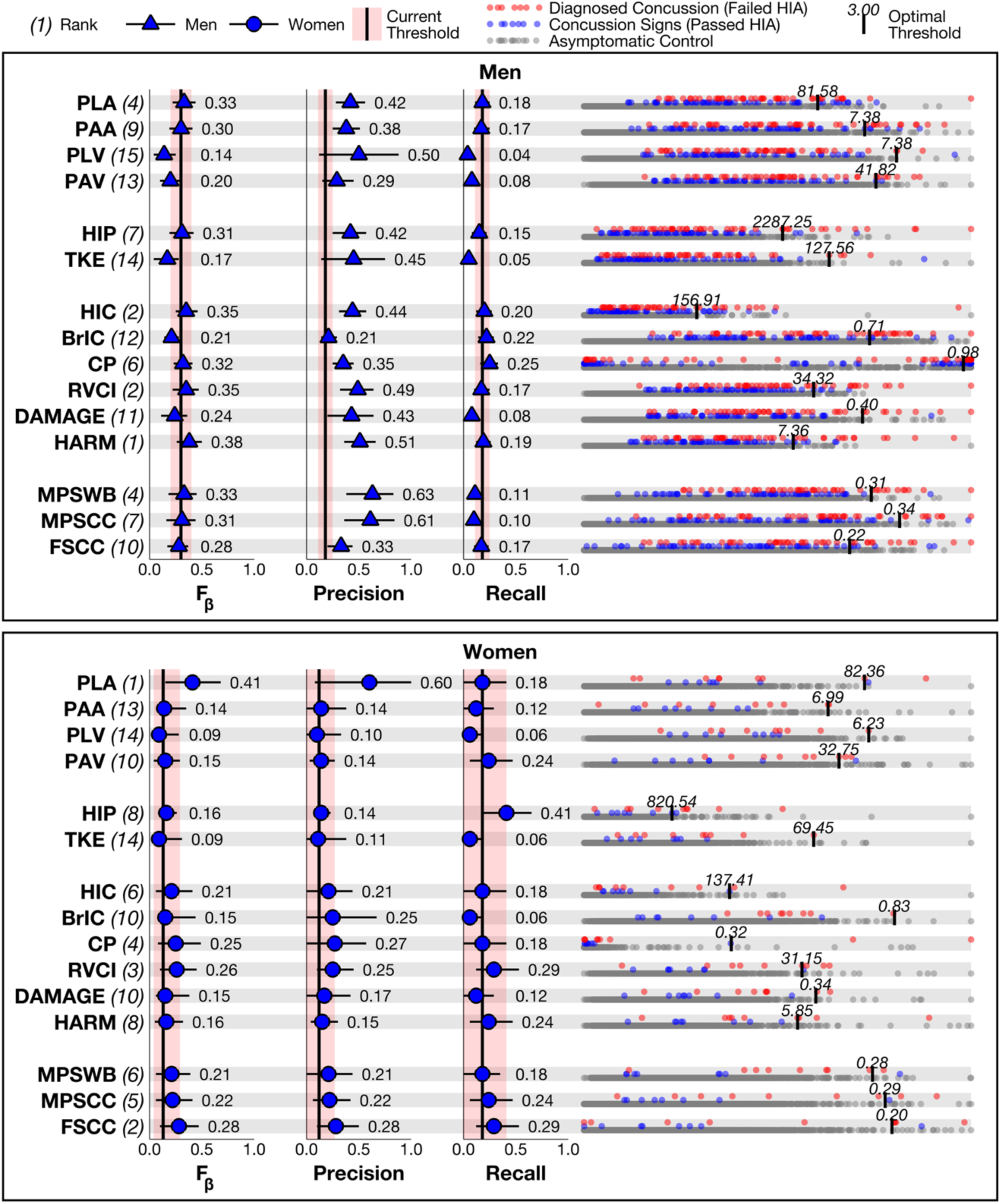
Screening performance of each severity metric for men and women based on PR analysis. Optimal decisioning thresholds were based on maximising F_β_ and rank, precision, and recall values correspond to those thresholds. Supplementary Tables 5 shows tabulated results. PLA = peak linear acceleration, PAA = peak angular acceleration, PLV = peak linear velocity, PAV = peak angular velocity, HIP = Head Impact Power, TKE = total kinetic energy, HIC = Head Injury Criterion, BrIC = Brain Injury Criterion, RVCI = Rotational Velocity Change Index, CP = Combined Probability, DAMAGE = Diffuse Axonal Multi-Axis General Evaluation, HARM = Head Acceleration Response Metric, MPS = maximum principal strain of the whole brain, MPSCC = maximum principal strain of the corpus callosum, FSCC = fibre strain of the corpus callosum

## Discussion

This study assesses the biomechanical severity of HAEs resulting in diagnosed concussion and observable concussion signs in elite men’s and women’s rugby league using a range of iMG-derived severity metrics, representing the largest iMG-measured concussion dataset reported to date. Diagnosed concussions had greater severity, on average, than asymptomatic HAEs across all metrics in both sexes, and sex differences in mean severity were observed for some metrics. Several iMG-derived severity metrics demonstrated meaningful diagnostic accuracy and yielded modest theoretical improvements over current iMG screening thresholds, supporting their potential role as objective adjuncts within the existing, clinician-led HIA pathway.

Group-level differences between diagnosed concussions and asymptomatic HAEs show that iMG-derived severity metrics captured clinically relevant differences in severity, but considerable overlap in their distributions indicates additional factors are involved. More advanced severity metrics may better approximate TBI mechanisms, however intrinsic factors mean a universal threshold for concussion is likely to remain elusive^31^. Biomechanical tolerance may be influenced by concussion history, with prior concussion increasing the risk of subsequent concussion^32^, and by prior HAE exposure within and before the match; in a case-control analysis, concussion cases were preceded by periods of greater HAE exposure than non-concussed controls matched by HAE magnitude^33^. Tolerance may also vary due to genetic variation and pre-existing neurological or psychiatric comorbidities^34,35^.

Furthermore, ‘concussion’ has been criticised as a diagnostic label describing a heterogeneous clinical syndrome rather than a single pathological entity^36^, therefore concussions may reflect distinct TBI mechanisms and symptom profiles that are not optimally captured by the same biomechanical severity metric.

Sex differences in concussion tolerance have been hypothesised due to brain morphology, particularly in axons, where males exhibit greater cross-sectional area and microtubule density^37^. These morphological differences have been shown to contribute to greater brain strains in women under the same loading conditions in finite-element simulations^38^. While higher average severity in men’s concussions in some severity metrics tentatively support hypothesised sex differences in concussion tolerance, a larger dataset of iMG-measured concussions in women is needed to investigate differences directly. Consequently, future research should continue to capture women’s concussions with iMGs.

Despite overlap in severity between outcomes, the diagnostic accuracy of iMG-derived severity metrics was comparable to that of existing HIA sub-tests^12^. When predicting clinical diagnosis, current HIA sub-tests (Maddocks questions, immediate memory, digits backwards, tandem gait, delayed recall) had ROC-AUC values of 0.60 or lower, while self-reported symptoms and clinical signs were both reported at 0.70^12^. The highest-performing iMG-derived severity metrics for clinical diagnosis in this study were RVCI in men and BrIC in women, with respective ROC-AUC values of 0.72 (CI:0.61-0.81) and 0.86 (CI:0.61-1.00). These findings support using iMG-derived severity metrics as an additional sub-test within the HIA, but do not support any single severity metric as a stand-alone diagnostic tool. Therefore, final concussion diagnosis should remain a clinician-led decision.

When considered as screening tools, optimal decision thresholds for several iMG-derived severity metrics theoretically improved performance compared to current thresholds. In men, the highest-performing severity metric was HARM, for which an optimal threshold of 7.36 increased precision by 0.16 and recall by 0.01 relative to current thresholds. In women, a PLA threshold of 82.36 *g* improved precision by 0.48 while maintaining recall. Based on the current sample across two seasons (27.0% of men’s player-hours and 31.5% of women’s player-hours), these optimal thresholds would theoretically false-positive screening alerts from 35 to 19 in men and 21 to 2 in women. However, recall remained below 0.20, meaning more than 80% of HAEs resulting in visible concussions signs failed to exceed these thresholds. Screening alert recall will remain constrained by the need for high precision to avoid excessive false-positive alerts overloading the HIA process^17^, therefore, multimodal approaches of identifying potential concussions are essential, including video replays and ongoing education to promote symptom self-reporting.

Beyond predictive performance, the feasibility of implementing iMG-derived severity metrics within the HIA depends on their computational burden and robustness to measurement error. More complex severity metrics requiring full time-series kinematic processing may be better suited to post-event diagnostic support than real-time screening alerts, where processing latency must remain low to allow timely removal from play. Currently, latency is minimised by processing data on-board iMGs, which may not be feasible with more advanced metrics. Metric stability under on-field noise is critical, as noise contamination can artificially inflate severity values and degrade predictive performance. Metrics derived from angular velocity are likely more stable than those based on angular or linear acceleration because angular velocity is measured directly by the gyroscope, whereas angular acceleration and linear acceleration at the head centre-of-gravity are derived from other signals via processes that amplify noise^39^.

### Clinical and Policy Implications

Given that iMG-derived severity metrics demonstrated diagnostic accuracy comparable to established HIA sub-tests^12^, incorporating iMG data as an additional sub-test in series with existing sub-tests to inform concussion diagnosis may be beneficial. For initiating off-field concussion assessment, several iMG-derived severity metrics provided modest theoretical improvements over current thresholds with greater precision, suggesting that current screening can be revised, however, any new implementation should be carefully monitored. Despite promising findings, current evidence does not support the use of iMGs as a stand-alone solution for diagnostics or screening; concussion diagnosis should remain a clinician-led decision and screening should continue to leverage multimodal approaches including visible concussion signs, sideline recognition, video review, and self-reporting by players.

### Limitations

Despite efforts to remove HAEs associated with visible concussion signs, the control pool may contained undiagnosed concussions, as concussion symptoms may be delayed and undetectable by video analysis alone^6^. Future research should evaluate the ability of iMG-derived severity metrics to detect concussions that do not present with visible signs.

Interpretation of these findings should consider the specific concussion detection and diagnosis processes used in these RFL competitions, as differences in how concussions are screened and diagnosed in other sports and settings may influence which HAEs are identified as concussive or non-concussive. The dataset is also not fully representative of all on-field data because HAEs with substantial noise were excluded, resulting in selection bias. Lastly, the women’s sample was relatively small and underpowered for sex-specific analyses, limiting inferences about women players.

## Conclusion

This study assessed the severity of HAEs resulting in diagnosed concussions in elite men’s and women’s rugby league using a range of biomechanical severity metrics, representing the largest dataset of iMG-measured concussions in any sport to date. Statistical comparisons showed that diagnosed concussions had, on average, greater severity than asymptomatic HAEs across all severity metrics in both sexes, with sex differences present in some metrics. The predictive capabilities of each severity metric were quantified, providing a basis for evaluating the diagnostic and screening utility of iMG-derived severity metrics within existing rugby HIA processes. Overall, these findings support integrating iMG-derived severity metrics into a multimodal, clinician-led HIA pathway as objective adjuncts for diagnosis and screening, while reinforcing that they cannot replace clinical judgement or other assessment modalities.

## Data Availability

All data produced in the present study are available upon reasonable request to the authors

## Statements

### Contributorship

JT, LW, SR, and BJ contributed to the conceptualisation and design of the study; JT, CO, SW, SS, DV, and BJ contributed to data acquisition; JT, MK, CZ, and SJ contributed to data processing and analysis; all authors contributed to interpretation of the results and to drafting and revising the manuscript for important intellectual content. All authors approved the final version.

### Data Sharing

All data relevant to the study are included in the article or uploaded as supplementary information.

### Ethics Approval

Ethical approval was granted by the university ethics committee and consent was obtained directly from participants.

### Patient Involvement

Neither patients nor members of the public were involved in planning, designing, collecting data, analysing and interpreting the results of this study.

### Equity, Diversity and Inclusions Statement

This study included players from across all teams participating in the Men’s and Women’s Super League competitions, inclusive of all races and ethnicities. Eleven of 16 clubs are based in the top 30% of most deprived local authorities according to the English Indices of Deprivation, with five in the top 10%. The author team consisted of male and female researchers from the UK, Canada, US, and South Africa, spanning PhD fellows, research analysts, consultant physicians, and associate and full professors.

## Supplementary Materials

**Supplementary Table 1.** Values used in severity metric estimates.

| Severity Measure(s) | Specific Values |
| --- | --- |
| Total Kinetic Energy (TKE) and Head Impact Power (HIP) | $m = 4.1$ kg (men) and 3.2 kg (women)<br>$I$ in x-axis = 0.0181 kg·m<sup>2</sup> (men) and 0.0114 kg·m<sup>2</sup> (women)<br>$I$ in y-axis = 0.0203 kg·m<sup>2</sup> (men) and 0.0138 kg·m<sup>2</sup> (women)<br>$I$ in z-axis = 0.0160 kg·m<sup>2</sup> (men) and 0.0119 kg·m<sup>2</sup> (women) |
| Brain Injury Criterion (BrIC) | $\omega_{xcr} = 66.2$<br>$\omega_{xcr} = 59.1$<br>$\omega_{xcr} = 44.2$ |
| Combined Probability (CP) | $\beta_0 = -12.60663$ (men) and -9.34360 (women)<br>$B_1 = 0.11215$ (men) and 0.08065 (women)<br>$\beta_2 = 1.08524$ (men) and 0.41566 (women)<br>$\beta_3 = -0.01024$ (men) and -0.00240 (women) |
| Rotational Velocity Change Index (RVCI) | $R_x = 1.00$<br>$R_y = 1.00$<br>$R_z = 1.17$ |

**Suplementary Figure 1.**
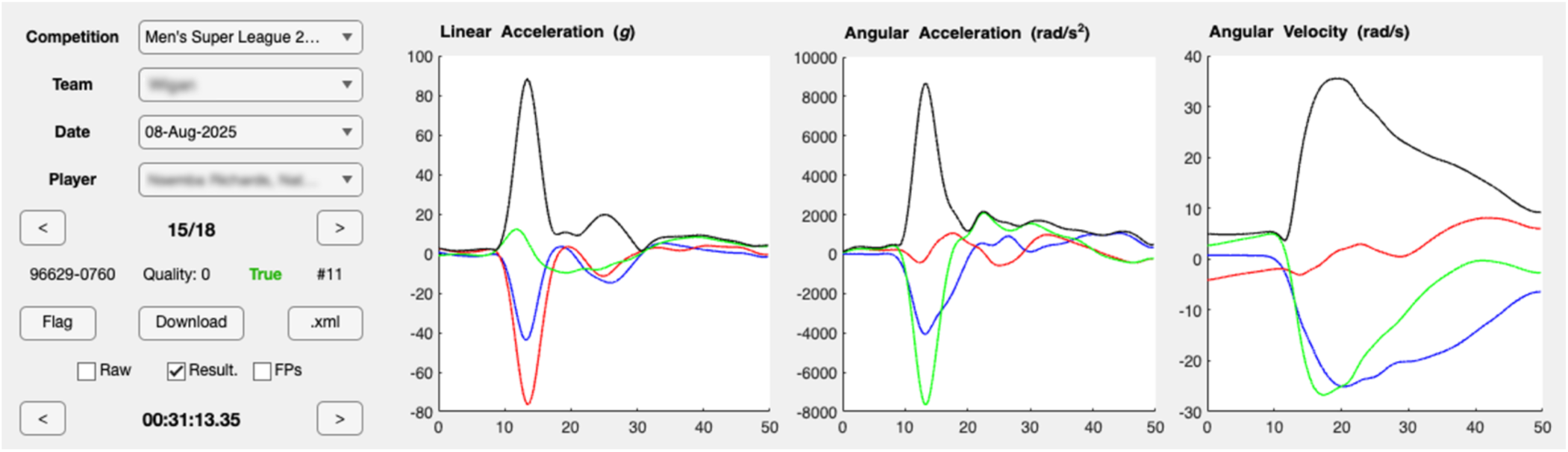
Graphical user interface used for video analysis of time-series iMG data in stage 3 of the matching and data quality screening procedure.

**Supplementary Table 2.** Mean (95% CI) values for each HAE outcome with statistical comparisons.

|  | Diagnosed SRC |  | Passed HIA |  | Asymptomatic Control |  |
| --- | --- | --- | --- | --- | --- | --- |
|  | Men | Women | Men | Women | Men | Women |
| *Peak Kinematics* |  |  |  |  |  |  |
| **PLA** | 59.66<br>(51.78-68.75) | 44.95<br>(32.84-61.52) | 49.37<br>(43.81-55.63) | 43.36<br>(31.08-60.49) | 19.17<br>(18.95-19.39) | 17.79<br>(17.42-18.17) |
| **PAA** | 5.28<br>(4.59-6.07) | 3.83<br>(2.81-5.22) | 4.28<br>(3.80-4.82) | 3.01<br>(2.17-4.19) | 1.69<br>(1.67-1.71) | 1.71<br>(1.68-1.75) |
| **PLV** | 4.02<br>(3.48-4.65) | 3.78<br>(2.74-5.22) | 3.57<br>(3.16-4.04) | 3.32<br>(2.36-4.67) | 2.36<br>(2.32-2.39) | 2.29<br>(2.23-2.34) |
| **PAV** | 27.27<br>(23.88-31.15) | 26.21<br>(19.53-35.16) | 21.96<br>(19.64-24.56) | 15.94<br>(11.67-21.78) | 13.07<br>(12.93-13.22) | 12.45<br>(12.20-12.70) |
| *Mechanical Energy Measures* |  |  |  |  |  |  |
| **HIP** | 1330.21<br>(1076.40-1643.86) | 669.15<br>(418.90-1068.91) | 906.79<br>(758.71-1083.77) | 412.69<br>(251.08-678.34) | 231.90<br>(228.35-235.51) | 152.63<br>(148.18-157.22) |
| **TKE** | 46.26<br>(35.33-60.57) | 28.10<br>(15.47-51.02) | 34.94<br>(27.84-43.85) | 19.69<br>(10.45-37.09) | 15.03<br>(14.65-15.43) | 9.69<br>(9.29-10.12) |
| *Composite Injury Criteria* |  |  |  |  |  |  |
| **HIC** | 91.68<br>(69.72-120.55) | 46.03<br>(25.11-84.35) | 61.76<br>(49.04-77.78) | 39.06<br>(20.54-74.29) | 8.43<br>(8.25-8.62) | 7.18<br>(6.90-7.48) |
| **BrIC** | 0.55<br>(0.48-0.61) | 0.56<br>(0.43-0.73) | 0.45<br>(0.41-0.50) | 0.33<br>(0.25-0.44) | 0.29<br>(0.29-0.30) | 0.29<br>(0.29-0.30) |
| **RVCI** | 27.90<br>(25.06-31.07) | 23.59<br>(18.59-29.93) | 21.87<br>(19.98-23.94) | 16.10<br>(12.51-20.73) | 11.00<br>(10.90-11.10) | 10.78<br>(10.61-10.95) |
| **CP** | 0.88<br>(0.15-0.99) | 0.03<br>(0.00-0.06) | 0.43<br>(0.02-0.93) | 0.01<br>(0.01-0.02) | 0.00<br>(0.00-0.00) | 0.00<br>(0.00-0.00) |
| **DAMAGE** | 0.26<br>(0.23-0.30) | 0.24<br>(0.18-0.32) | 0.21<br>(0.19-0.23) | 0.16<br>(0.12-0.21) | 0.12<br>(0.12-0.12) | 0.11<br>(0.11-0.12) |
| **HARM** | 5.65<br>(4.98-6.40) | 4.69<br>(3.56-6.19) | 4.37<br>(3.94-4.86) | 3.16<br>(2.35-4.24) | 2.03<br>(2.01-2.05) | 1.92<br>(1.88-1.96) |
| *Brain Strain Metrics* |  |  |  |  |  |  |
| **MPS** | 0.25<br>(0.23-0.27) | 0.21<br>(0.17-0.25) | 0.20<br>(0.19-0.22) | 0.14<br>(0.11-0.17) | 0.13<br>(0.13-0.13) | 0.13<br>(0.13-0.13) |
| **MPSCC** | 0.25<br>(0.23-0.28) | 0.20<br>(0.17-0.24) | 0.21<br>(0.20-0.23) | 0.14<br>(0.11-0.17) | 0.13<br>(0.13-0.13) | 0.12<br>(0.12-0.13) |
| **FSCC** | 0.17<br>(0.16-0.19) | 0.12<br>(0.10-0.15) | 0.14<br>(0.13-0.15) | 0.09<br>(0.07-0.11) | 0.09<br>(0.09-0.09) | 0.08<br>(0.08-0.08) |

PLA = peak linear acceleration, PAA = peak angular acceleration, PLV = peak linear velocity, PAV = peak angular velocity, HIP = Head Impact Power, TKE = total kinetic energy, HIC = Head Injury Criterion, BrIC = Brain Injury Criterion, RVCI = Rotational Velocity Change Index, CP = Combined Probability, DAMAGE = Diffuse Axonal Multi-Axis General Evaluation, HARM = Head Acceleration Response Metric, MPS = maximum principal strain of the whole brain, MPSCC = maximum principal strain of the corpus callosum, FSCC = fibre strain of the corpus callosum, IQR = interquartile range, CI = confidence interval

**Supplementary Table 3.** Pairwise comparisons within each sex between HAE outcomes.

|  | Diagnosed Concussion – Concussion Signs |  | Diagnosed Concussion - Asymptomatic Control |  | Concussion Signs - Asymptomatic Control |  |
| --- | --- | --- | --- | --- | --- | --- |
|  | Estimated Difference (%) | p-value | Estimated Difference (%) | p-value | Estimated Difference (%) | p-value |
| **Men** |  |  |  |  |  |  |
| *Peak Kinematics* |  |  |  |  |  |  |
| **PLA** | 20.85% | 0.11 | 211.28% | < 0.05 | 157.57% | < 0.05 |
| **PAA** | 23.31% | 0.06 | 212.90% | < 0.05 | 153.75% | < 0.05 |
| **PLV** | 12.70% | 0.43 | 70.68% | < 0.05 | 51.44% | < 0.05 |
| **PAV** | 24.20% | < 0.05 | 108.60% | < 0.05 | 67.96% | < 0.05 |
| *Mechanical Energy Measures* |  |  |  |  |  |  |
| **HIP** | 46.69% | < 0.05 | 473.60% | < 0.05 | 291.02% | < 0.05 |
| **TKE** | 32.40% | 0.26 | 207.71% | < 0.05 | 132.40% | < 0.05 |
| *Composite Injury Criteria* |  |  |  |  |  |  |
| **HIC** | 48.44% | 0.08 | 987.02% | < 0.05 | 632.30% | < 0.05 |
| **BrIC** | 20.75% | < 0.05 | 85.38% | < 0.05 | 53.52% | < 0.05 |
| **CP** | n/a | n/a | n/a | n/a | n/a | n/a |
| **RVCI** | 27.60% | < 0.05 | 153.72% | < 0.05 | 98.84% | < 0.05 |
| **DAMAGE** | 27.26% | < 0.05 | 121.01% | < 0.05 | 73.68% | < 0.05 |
| **HARM** | 29.11% | < 0.05 | 177.84% | < 0.05 | 115.19% | < 0.05 |
| *Brain Strain Metrics* |  |  |  |  |  |  |
| **MPS** | 21.40% | < 0.05 | 86.20% | < 0.05 | 53.38% | < 0.05 |
| **MPSCC** | 19.69% | < 0.05 | 93.06% | < 0.05 | 61.30% | < 0.05 |
| **FSCC** | 22.41% | < 0.05 | 99.48% | < 0.05 | 62.96% | < 0.05 |
| **Women** |  |  |  |  |  |  |
| *Peak Kinematics* |  |  |  |  |  |  |
| **PLA** | 3.67% | 0.99 | 152.62% | < 0.05 | 143.67% | < 0.05 |
| **PAA** | 26.90% | 0.56 | 123.10% | < 0.05 | 75.80% | < 0.05 |
| **PLV** | 13.91% | 0.85 | 65.57% | < 0.05 | 45.35% | 0.08 |
| **PAV** | 64.41% | 0.06 | 110.52% | < 0.05 | 28.05% | 0.27 |
| *Mechanical Energy Measures* |  |  |  |  |  |  |
| **HIP** | 62.14% | 0.35 | 338.40% | < 0.05 | 170.38% | < 0.05 |
| **TKE** | 42.71% | 0.7 | 189.84% | < 0.05 | 103.10% | 0.07 |
| *Composite Injury Criteria* |  |  |  |  |  |  |
| **HIC** | 17.83% | 0.93 | 540.73% | < 0.05 | 443.75% | < 0.05 |
| **BrIC** | 71.05% | < 0.05 | 90.79% | < 0.05 | 11.54% | 0.73 |
| **CP** | n/a | n/a | n/a | n/a | n/a | n/a |
| **RVCI** | 46.48% | 0.08 | 118.81% | < 0.05 | 49.38% | < 0.05 |
| **DAMAGE** | 49.78% | 0.11 | 111.34% | < 0.05 | 41.09% | 0.05 |
| **HARM** | 48.62% | 0.13 | 144.60% | < 0.05 | 64.58% | < 0.05 |
| *Brain Strain Metrics* |  |  |  |  |  |  |
| **MPS** | 50.37% | < 0.05 | 63.27% | < 0.05 | 8.58% | 0.7 |
| **MPSCC** | 44.69% | < 0.05 | 61.27% | < 0.05 | 11.46% | 0.55 |
| **FSCC** | 43.38% | 0.08 | 52.43% | < 0.05 | 6.32% | 0.87 |

PLA = peak linear acceleration, PAA = peak angular acceleration, PLV = peak linear velocity, PAV = peak angular velocity, HIP = Head Impact Power, TKE = total kinetic energy, HIC = Head Injury Criterion, BrIC = Brain Injury Criterion, RVCI = Rotational Velocity Change Index, CP = Combined Probability, DAMAGE = Diffuse Axonal Multi-Axis General Evaluation, HARM = Head Acceleration Response Metric, MPS = maximum principal strain of the whole brain, MPSCC = maximum principal strain of the corpus callosum, FSCC = fibre strain of the corpus callosum, IQR = interquartile range

**Supplementary Table 4.**
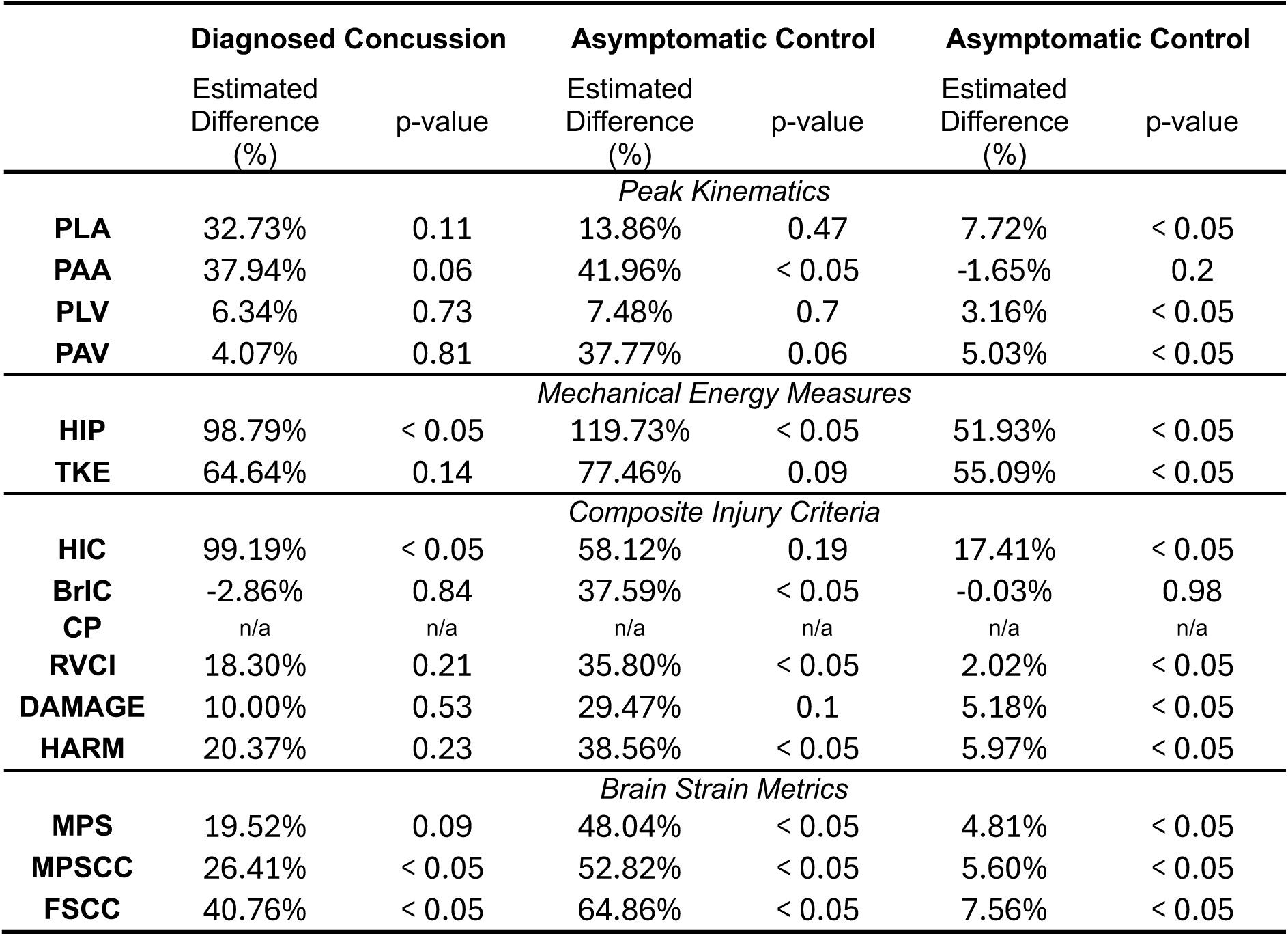
Pairwise comparisons between sexes for each HAE outcome.

**Supplementary Table 5.** Diagnostic accuracy (i.e., prediction 1) of each severity metric for men and women. Optimal decisioning threshold were based on *J* values and TPR and FPR values correspond to those thresholds.

|  | Men |  |  |  |  |  | Women |  |  |  |  |  |
| --- | --- | --- | --- | --- | --- | --- | --- | --- | --- | --- | --- | --- |
|  | ROC-AUC | Rank | Thres-hold | *J* | TPR | FPR | ROC-AUC | Rank | Thres-hold | *J* | TPR | FPR |
| *Peak Kinematics* |  |  |  |  |  |  |  |  |  |  |  |  |
| **PLA** | 0.63<br>(0.52-0.73) | 13 | 55.06 | 0.28<br>(0.09-0.46) | 0.64<br>(0.48-0.77) | 0.35<br>(0.24-0.48) | 0.61<br>(0.32-0.89) | 14 | 42.51 | 0.42<br>(-0.08-0.78) | 0.67<br>(0.33-1.00) | 0.25<br>(0.00-0.62) |
| **PAA** | 0.65<br>(0.55-0.75) | 10 | 5.66 | 0.24<br>(0.07-0.43) | 0.50<br>(0.34-0.66) | 0.26<br>(0.16-0.37) | 0.74<br>(0.46-0.96) | 9 | 5.04 | 0.56<br>(0.00-0.89) | 0.56<br>(0.22-0.89) | 0.00<br>(0.00-0.31) |
| **PLV** | 0.61<br>(0.49-0.71) | 15 | 3.88 | 0.30<br>(0.11-0.48) | 0.66<br>(0.52-0.80) | 0.35<br>(0.26-0.47) | 0.71<br>(0.42-0.93) | 10 | 4.38 | 0.44<br>(-0.03-0.78) | 0.44<br>(0.06-0.78) | 0.00<br>(0.00-0.38) |
| **PAV** | 0.69<br>(0.57-0.78) | 3 | 23.81 | 0.32<br>(0.12-0.49) | 0.73<br>(0.59-0.86) | 0.40<br>(0.29-0.53) | 0.83<br>(0.55-1.00) | 2 | 23.94 | 0.65<br>(0.17-1.00) | 0.78<br>(0.56-1.00) | 0.12<br>(0.00-0.50) |
| *Mechanical Energy Measures* |  |  |  |  |  |  |  |  |  |  |  |  |
| **HIP** | 0.67<br>(0.56-0.77) | 6 | 1702.5<br>4 | 0.28<br>(0.11-0.44) | 0.41<br>(0.25-0.55) | 0.13<br>(0.06-0.23) | 0.78<br>(0.50-0.97) | 6 | 924.39 | 0.56<br>(0.10-0.89) | 0.56<br>(0.22-0.89) | 0.00<br>(0.00-0.00) |
| **TKE** | 0.63<br>(0.52-0.73) | 13 | 44.7 | 0.31<br>(0.12-0.48) | 0.64<br>(0.50-0.77) | 0.32<br>(0.23-0.45) | 0.71<br>(0.40-0.92) | 10 | 36.08 | 0.43<br>(-0.01-0.78) | 0.56<br>(0.22-0.89) | 0.12<br>(0.00-0.38) |
| *Composite Injury Criteria* |  |  |  |  |  |  |  |  |  |  |  |  |
| **HIC** | 0.64<br>(0.53-0.73) | 12 | 69.47 | 0.26<br>(0.09-0.43) | 0.68<br>(0.55-0.82) | 0.42<br>(0.31-0.53) | 0.53<br>(0.24-0.83) | 15 | 82.46 | 0.32<br>(-0.17-0.67) | 0.44<br>(0.11-0.78) | 0.12<br>(0.00-0.50) |
| **BrIC** | 0.68<br>(0.56-0.77) | 4 | 0.58 | 0.30<br>(0.12-0.47) | 0.55<br>(0.39-0.70) | 0.24<br>(0.15-0.35) | 0.86<br>(0.61-1.00) | 1 | 0.42 | 0.75<br>(0.38-1.00) | 1.00<br>(1.00-1.00) | 0.25<br>(0.00-0.62) |
| **RVCI** | 0.65<br>(0.54-0.75) | 1 | 0.71 | 0.25<br>(0.07-0.43) | 0.59<br>(0.45-0.73) | 0.34<br>(0.23-0.45) | 0.62<br>(0.32-0.90) | 2 | 0.03 | 0.54<br>(0.08-0.89) | 0.67<br>(0.33-1.00) | 0.12<br>(0.00-0.38) |
| **CP** | 0.72<br>(0.61-0.81) | 10 | 27.61 | 0.32<br>(0.13-0.51) | 0.55<br>(0.41-0.69) | 0.23<br>(0.13-0.34) | 0.83<br>(0.56-1.00) | 13 | 18.93 | 0.76<br>(0.28-1.00) | 0.89<br>(0.67-1.00) | 0.12<br>(0.00-0.50) |
| **DAMAGE** | 0.67<br>(0.57-0.77) | 6 | 0.22 | 0.27<br>(0.07-0.43) | 0.75<br>(0.61-0.86) | 0.48<br>(0.37-0.61) | 0.81<br>(0.56-1.00) | 4 | 0.23 | 0.65<br>(0.19-1.00) | 0.78<br>(0.44-1.00) | 0.12<br>(0.00-0.38) |
| **HARM** | 0.68<br>(0.56-0.77) | 4 | 4.76 | 0.29<br>(0.11-0.46) | 0.73<br>(0.59-0.84) | 0.44<br>(0.31-0.56) | 0.75<br>(0.47-0.96) | 8 | 3.98 | 0.51<br>(0.12-0.88) | 0.89<br>(0.67-1.00) | 0.38<br>(0.12-0.75) |
| *Brain Strain Metrics* |  |  |  |  |  |  |  |  |  |  |  |  |
| **MPS** | 0.70<br>(0.59-0.79) | 2 | 0.26 | 0.30<br>(0.12-0.48) | 0.48<br>(0.34-0.61) | 0.18<br>(0.10-0.29) | 0.81<br>(0.54-1.00) | 4 | 0.18 | 0.76<br>(0.39-1.00) | 0.89<br>(0.67-1.00) | 0.12<br>(0.00-0.50) |
| **MPSCC** | 0.67<br>(0.57-0.77) | 6 | 0.28 | 0.28<br>(0.09-0.46) | 0.48<br>(0.32-0.61) | 0.19<br>(0.11-0.31) | 0.78<br>(0.50-0.99) | 6 | 0.18 | 0.54<br>(0.07-0.89) | 0.67<br>(0.44-1.00) | 0.12<br>(0.00-0.50) |
| **FSCC** | 0.67<br>(0.56-0.77) | 6 | 0.17 | 0.35<br>(0.15-0.54) | 0.66<br>(0.51-0.80) | 0.31<br>(0.19-0.42) | 0.68<br>(0.39-0.93) | 12 | 0.11 | 0.54<br>(-0.04-0.89) | 0.67<br>(0.33-1.00) | 0.12<br>(0.00-0.50) |

ROC-AUC = area under the ROC curve, *J* = Youden's Index, TPR = true positive rate, FPR = false positive rate, PLA = peak linear acceleration, PAA = peak angular acceleration, PLV = peak linear velocity, PAV = peak angular velocity, HIP = Head Impact Power, TKE = total kinetic energy, HIC = Head Injury Criterion, BrIC = Brain Injury Criterion, RVCI = Rotational Velocity Change Index, CP = Combined Probability, DAMAGE = Diffuse Axonal Multi-Axis General Evaluation, HARM = Head Acceleration Response Metric, MPS = maximum principal strain of the whole brain, MPSCC = maximum principal strain of the corpus callosum, FSCC = fibre strain of the corpus callosum, HAE = head acceleration event

**Supplementary Table 5.** Screening accuracy (i.e., prediction 2) of each severity metric for men and women. Optimal decisioning threshold were based on *J* values and TPR and FPR values correspond to those thresholds.

|  | Men |  |  |  |  | Women |  |  |  |  |
| --- | --- | --- | --- | --- | --- | --- | --- | --- | --- | --- |
|  | Thres-<br>hold | F<sub>β</sub> | Rank | Precision | Recall | Thres-<br>hold | F<sub>β</sub> | Rank | Precision | Recall |
| **Current Threshold** | n/a | 0.30<br>(0.20-0.40) | n/a | 0.35 (0.24-0.47) | 0.18<br>(0.11-0.25) | n/a | 0.13<br>(0.04-0.29) | n/a | 0.12 (0.00-0.27) | 0.18<br>(0.00-0.41) |
| *Peak Kinematics* |  |  |  |  |  |  |  |  |  |  |
| **PLA** | 81.58 | 0.33<br>(0.22-0.44) | 4 | 0.42<br>(0.28-0.56) | 0.18<br>(0.11-0.25) | 82.36 | 0.41<br>(0.15-0.68) | 1 | 0.60<br>(0.08-1.00) | 0.18<br>(0.00-0.41) |
| **PAA** | 7.38 | 0.30<br>(0.19-0.41) | 9 | 0.38<br>(0.25-0.51) | 0.17<br>(0.10-0.25) | 6.99 | 0.14<br>(0.06-0.35) | 13 | 0.14<br>(0.00-0.38) | 0.12<br>(0.00-0.29) |
| **PLV** | 7.38 | 0.14<br>(0.04-0.25) | 15 | 0.50<br>(0.12-0.88) | 0.04<br>(0.01-0.08) | 6.23 | 0.09<br>(0.06-0.28) | 14 | 0.10<br>(0.00-0.33) | 0.06<br>(0.00-0.18) |
| **PAV** | 41.82 | 0.20<br>(0.10-0.31) | 13 | 0.29<br>(0.15-0.45) | 0.08<br>(0.04-0.14) | 32.75 | 0.15<br>(0.04-0.29) | 10 | 0.14<br>(0.03-0.27) | 0.24<br>(0.06-0.47) |
| *Mechanical Energy Measures* |  |  |  |  |  |  |  |  |  |  |
| **HIP** | 2287.25 | 0.31<br>(0.19-0.42) | 7 | 0.42<br>(0.25-0.57) | 0.15<br>(0.08-0.22) | 820.54 | 0.16<br>(0.08-0.26) | 8 | 0.14<br>(0.07-0.23) | 0.41<br>(0.18-0.65) |
| **TKE** | 127.56 | 0.17<br>(0.04-0.28) | 14 | 0.45<br>(0.14-0.75) | 0.05<br>(0.01-0.08) | 69.45 | 0.09<br>(0.07-0.31) | 14 | 0.11<br>(0.00-0.38) | 0.06<br>(0.00-0.18) |
| *Composite Injury Criteria* |  |  |  |  |  |  |  |  |  |  |
| **HIC** | 156.91 | 0.35<br>(0.25-0.46) | 2 | 0.44<br>(0.31-0.57) | 0.20<br>(0.12-0.27) | 137.41 | 0.21<br>(0.06-0.41) | 6 | 0.21<br>(0.00-0.44) | 0.18<br>(0.00-0.41) |
| **BrIC** | 0.71 | 0.21<br>(0.14-0.28) | 12 | 0.21<br>(0.14-0.29) | 0.22<br>(0.14-0.29) | 0.83 | 0.15<br>(0.11-0.44) | 10 | 0.25<br>(0.00-0.67) | 0.06<br>(0.00-0.18) |
| **RVCI** | 0.98 | 0.32<br>(0.23-0.41) | 2 | 0.35<br>(0.25-0.45) | 0.25<br>(0.16-0.33) | 0.32 | 0.25<br>(0.08-0.49) | 4 | 0.27<br>(0.00-0.57) | 0.18<br>(0.00-0.41) |
| **CP** | 34.32 | 0.35<br>(0.22-0.47) | 6 | 0.49<br>(0.32-0.64) | 0.17<br>(0.10-0.25) | 31.15 | 0.26<br>(0.10-0.45) | 3 | 0.25<br>(0.10-0.45) | 0.29<br>(0.12-0.53) |
| **DAMAGE** | 0.4 | 0.24<br>(0.11-0.36) | 11 | 0.43<br>(0.20-0.64) | 0.08<br>(0.04-0.14) | 0.34 | 0.15<br>(0.06-0.38) | 10 | 0.17<br>(0.00-0.42) | 0.12<br>(0.00-0.29) |
| **HARM** | 7.36 | 0.38<br>(0.26-0.50) | 1 | 0.51<br>(0.36-0.66) | 0.19<br>(0.12-0.26) | 5.85 | 0.16<br>(0.05-0.32) | 8 | 0.15<br>(0.04-0.30) | 0.24<br>(0.06-0.47) |
| *Brain Strain Metrics* |  |  |  |  |  |  |  |  |  |  |
| **MPS** | 0.31 | 0.33<br>(0.18-0.45) | 4 | 0.63<br>(0.38-0.83) | 0.11<br>(0.06-0.17) | 0.28 | 0.21<br>(0.06-0.39) | 6 | 0.21<br>(0.00-0.44) | 0.18<br>(0.00-0.35) |
| **MPSCC** | 0.34 | 0.31<br>(0.16-0.44) | 7 | 0.61<br>(0.36-0.84) | 0.10<br>(0.05-0.16) | 0.29 | 0.22<br>(0.06-0.41) | 5 | 0.22<br>(0.06-0.42) | 0.24<br>(0.06-0.47) |
| **FSCC** | 0.22 | 0.28<br>(0.17-0.37) | 10 | 0.33<br>(0.20-0.44) | 0.17<br>(0.10-0.24) | 0.20 | 0.28<br>(0.10-0.47) | 2 | 0.28<br>(0.10-0.50) | 0.29<br>(0.12-0.53) |

PLA = peak linear acceleration, PAA = peak angular acceleration, PLV = peak linear velocity, PAV = peak angular velocity, HIP = Head Impact Power, TKE = total kinetic energy, HIC = Head Injury Criterion, BrIC = Brain Injury Criterion, RVCI = Rotational Velocity Change Index, CP = Combined Probability, DAMAGE = Diffuse Axonal Multi-Axis General Evaluation, HARM = Head Acceleration Response Metric, MPS = maximum principal strain of the whole brain, MPSCC = maximum principal strain of the corpus callosum, FSCC = fibre strain of the corpus callosum, HAE = head acceleration event

